# Recent and forecasted increases in coccidioidomycosis incidence in California linked to hydroclimatic swings

**DOI:** 10.1101/2024.08.30.24312858

**Authors:** Simon K. Camponuri, Alexandra K. Heaney, Gail Sondermeyer Cooksey, Duc J. Vugia, Seema Jain, Daniel L. Swain, John Balmes, Justin V. Remais, Jennifer R Head

## Abstract

Coccidioidomycosis, or Valley fever, is an infectious disease caused by inhalation of *Coccidioides* spp., fungi found primarily in soils of the southwestern United States. Prior work showed that coccidioidomycosis cases in California sharply increase by nearly 2-fold following wet winters that occur one- and two-years following drought. Statewide drought between 2020-2022 followed by heavy precipitation during the 2022-2023 winter raised concerns over potential increases in coccidioidomycosis cases in the fall of 2023, prompting California Department of Public Health (CDPH) to issue public health alerts. As anticipated, California saw a near record number of cases in 2023, with 9,054 provisional cases reported. During the 2023-2024 California wet season, precipitation was 115% the long-term average, furthering concerns about continued high coccidioidomycosis risk. We developed an ensemble model to forecast coccidioidomycosis cases in California in 2024-2025. Using this model, we predicted a total of 11,846 cases (90% PI: 10,056–14,094) in California between April 1, 2023, and March 31, 2024, encompassing the preliminary state report of 10,593. Our model forecasted 12,244 cases statewide between April 1, 2024, and March 31, 2025 — a 62% increase over the cases reported during the same period two years prior, and on par with the high incidence seen in 2023. The Southern San Joaquin Valley (5,398 cases, 90% PI: 4,556–6,442), Southern Coast (3,322, 90% PI: 2,694–3,961), and Central Coast (1,207 cases, 90% PI: 867–1,585) regions are expected to see the largest number of infections. Our model forecasts that disease incidence will exhibit pronounced seasonality, particularly in endemic regions, with cases rising in June and peaking in November at 1,411 (90% PI: 815–2,172) cases statewide – 98% higher than the peak two years prior (714) and nearly as high as the peak in 2023 (1,462). Near-term forecasts have the potential to inform public health messaging to enhance provider and patient awareness, encourage risk reduction practices, and improve recognition and management of coccidioidomycosis.

## Introduction

Coccidioidomycosis, or Valley fever, is an infectious disease caused by inhalation of *Coccidioides* spp., pathogenic fungi found primarily in soils of the southwestern United States. In 2023, California reported the second highest number of coccidioidomycosis cases on record, with 9,054 provisional cases reported statewide (only surpassed by 9,093 cases in 2019), following an already dramatic 8-fold rise in coccidioidomycosis incidence in California between 2000 and 2018 (1). The number of cases reported during the 2023 calendar year was 21% higher than the prior calendar year. In California, cases typically begin to rise in the late summer, peak in the late fall or early winter, and return to low levels by March (2). Between Apr. 2023 and Mar. 2024, a period of time capturing the full 2023 transmission season (2), California reported 10,593 cases, 40% higher than the same period the year prior (3).

The 2023 spike in incidence may be attributable, in part, to a swing from extreme drought to extreme precipitation that occurred in the winter of 2022-2023. Between early 2020 and late 2022, California endured severe drought, with 88% of the state experiencing extreme dryness (4). An unusually wet winter followed in 2022-2023, with statewide precipitation exceeding 150% of average, ranking it among the top 10 wettest seasons in the past century (4). Alternation between dry and wet years has been linked to increased coccidioidomycosis incidence in California (2, 5). Availability of soil moisture during wet winter seasons is hypothesized to support fungal growth, contributing to an abundance of spores available for airborne dispersal during hot, dry conditions characteristic of California summer and early fall (5). Drought preceding the rainy season may enhance fungal growth by eliminating microbial competitors from the soil (6) or by affecting rodent populations, a putative reservoir host and/or nutrient source for the fungus (7). Prior work found that high precipitation following the 2007-2009 and 2012-2015 droughts in California led to 1,467 and 2,649 excess coccidioidomycosis cases, respectively, in the two years following each drought (5).

Statewide precipitation during the 2023-2024 California wet season was 115% of the long-term average, marking the second consecutive wetter-than-average season following a severe drought (4). This suggests the possibility of continued high coccidioidomycosis incidence during the 2024 transmission year, with cases expected to rise in fall 2024 and continue into early 2025. Disease forecasts can guide the creation and dissemination of state and local public health alerts and messages by pinpointing when and where disease risk is expected to be highest. Such targeted public health messaging can raise provider and patient awareness about disease risk and presentation, leading to earlier diagnosis, enhanced patient monitoring, and more effective disease management, ultimately improving patient outcomes. As an example, following the wet winter of 2022-2023, California Department of Public Health (CDPH) issued a press release in August 2023 alerting the public and healthcare providers of potential increased Valley fever risk and providing exposure mitigation information (8). In January 2024, CDPH released a Health Advisory to healthcare facilities, recommending that they consider coccidioidomycosis in differential diagnoses of community acquired pneumonia (CAP) or other respiratory illnesses in endemic or emerging areas (9).

Here, we develop an ensemble prediction model to forecast cases of coccidioidomycosis across regions of California. We demonstrate accurate predictive ability of this model during the 2023 transmission year and use the model to develop region-specific monthly coccidioidomycosis forecasts for the 2024-2025 transmission year. These forecasts can aid the state’s public health officials, physicians, state planners, and residents in preparing for, preventing, and responding to potential increased infection risk.

## Methods

To develop a predictive algorithm for generating short-term disease forecasts, we adapted our ensemble modeling approach published previously (5). In brief, we generated five candidate prediction algorithms: four generalized linear models with varying interactions and covariates, and one random forest model. Eligible predictors included season, year, soil texture, elevation, percent impervious surface, lagged total monthly precipitation, and lagged mean temperature (Table S1). We then examined how each candidate algorithm performed when used to generate short-term out-of-sample predictions by using a progressive time-series cross validation approach to determine the out-of-sample future prediction error of each model, examining model performance in time periods after the training data (Fig. 1) (10). Ensemble forecasts were weighted averages of individual model forecasts, with weights proportional to the inverse of their out-of-sample prediction error. We fit separate ensemble models for each county (for highly endemic regions) or region (for low to moderately endemic regions; Fig. S1) in California relating monthly reported cases per census tract to climatological or environmental predictors.

**Fig. 1.**
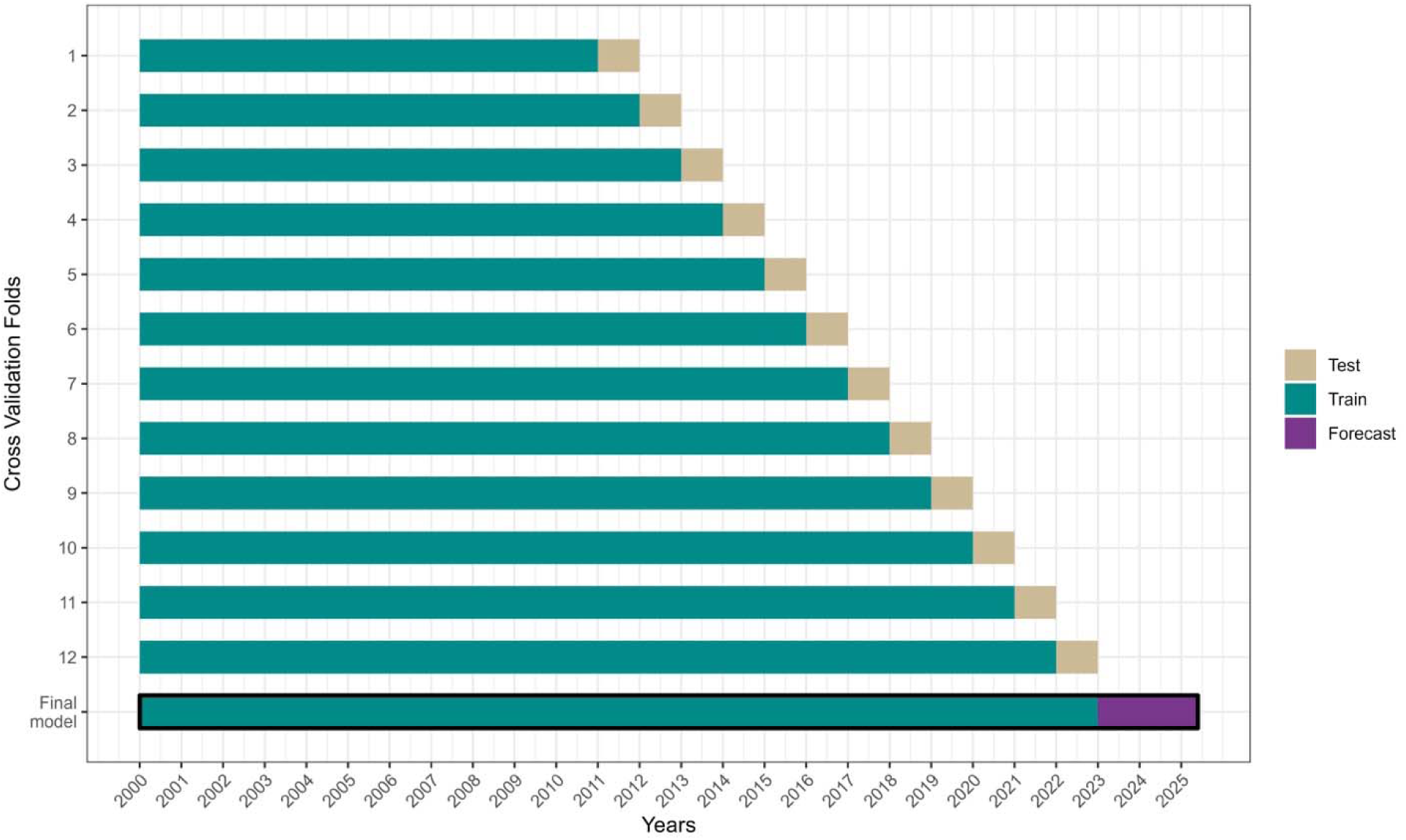
Schematic of the progressive time-series cross-validation approach used to generate weights for the ensemble model. Each progressive fold added an additional year to the training data (green) and predicted the following test year (beige). For example, we fit the models to data from 2000-2011 and predicted cases in 2012. We then fit the model to data from 2000-2012 and predicted cases in 2013, and so on. Individual models were then weighted by the inverse of their out-of-sample error across test years and used to forecast cases from Jan. 1^st^, 2023 – Mar. 31^st^, 2025 (purple).

To generate forecasts of coccidioidomycosis cases, we applied each model to temperature and precipitation data obtained from the PRISM Climate Group from January 2023 through March 2025 (11). We estimated temperature and precipitation beyond April 2024 (the date of analysis) by extrapolating monthly temperature averages in each census tract using a 42-year linear trend, and by setting precipitation equal to the 50^th^ percentile of the 42-year precipitation distribution (1981–2023). As the future climate is uncertain, we examined the sensitivity of forecasts to two alternative precipitation scenarios, drier-than-average (20^th^ percentile) and wetter-than-average (80^th^ percentile), and two alternative temperature scenarios, warmer-than-average (+3°F) and cooler-than-average (−3°F). We fit each model to census tract-level case data reported to CDPH from January 1, 2000, to December 31, 2022, and then took a weighted average (with weights as described above) of the fitted models to create an ensemble forecast of cases for each month between January 1, 2023, and March 31, 2025. We summed cases across calendar years and the coccidioidomycosis “transmission year” (2), which spans April 1^st^ to March 31^st^.

To quantify uncertainty in our forecasts, we generated 90% prediction intervals using a two-step bootstrapping process. We first sampled census tracts with replacement to generate 500 bootstrapped datasets, fit the models to each bootstrapped dataset, and predicted cases across the study period using each model. We then resampled each model’s predictions from a quasi-Poisson distribution with the mean set to each bootstrapped prediction and calculated the 5^th^ and 95^th^ percentiles of the resulting distributions (Fig. S2). Our 90% prediction intervals were conservative, capturing 91%–100% of observed cases. All analyses were conducted in R (version 4.3.2; R Core Team).

## Results

### Accurate predictions of high coccidioidomycosis cases during 2023-2024 season

Our ensemble model predicted a total of 11,846 cases (90% PI: 10,056–14,094) in California between April 1, 2023, and March 31, 2024, encompassing the preliminary state report of 10,593 (as of July 31, 2024; Fig. 2) (3). The model overpredicted cases during spring and summer months when incidence was low but more closely matched provisional cases during months with higher incidence (Fig. 2). Preliminary data show that the peak number of monthly cases in California was 1,462, which aligns with our model prediction of 1,619 (90% PI: 928–2,507). Regionally, our model predicted the greatest number of cases in the Southern San Joaquin Valley (5,557 cases, 90% PI: 4,778–6,757), Southern Coast (3,049 cases, 90% PI: 2,495–3,739), and Central Coast (1,189 cases, 90% PI: 819–1,599) regions (Fig. 3; Table 1).

**Fig. 2.**
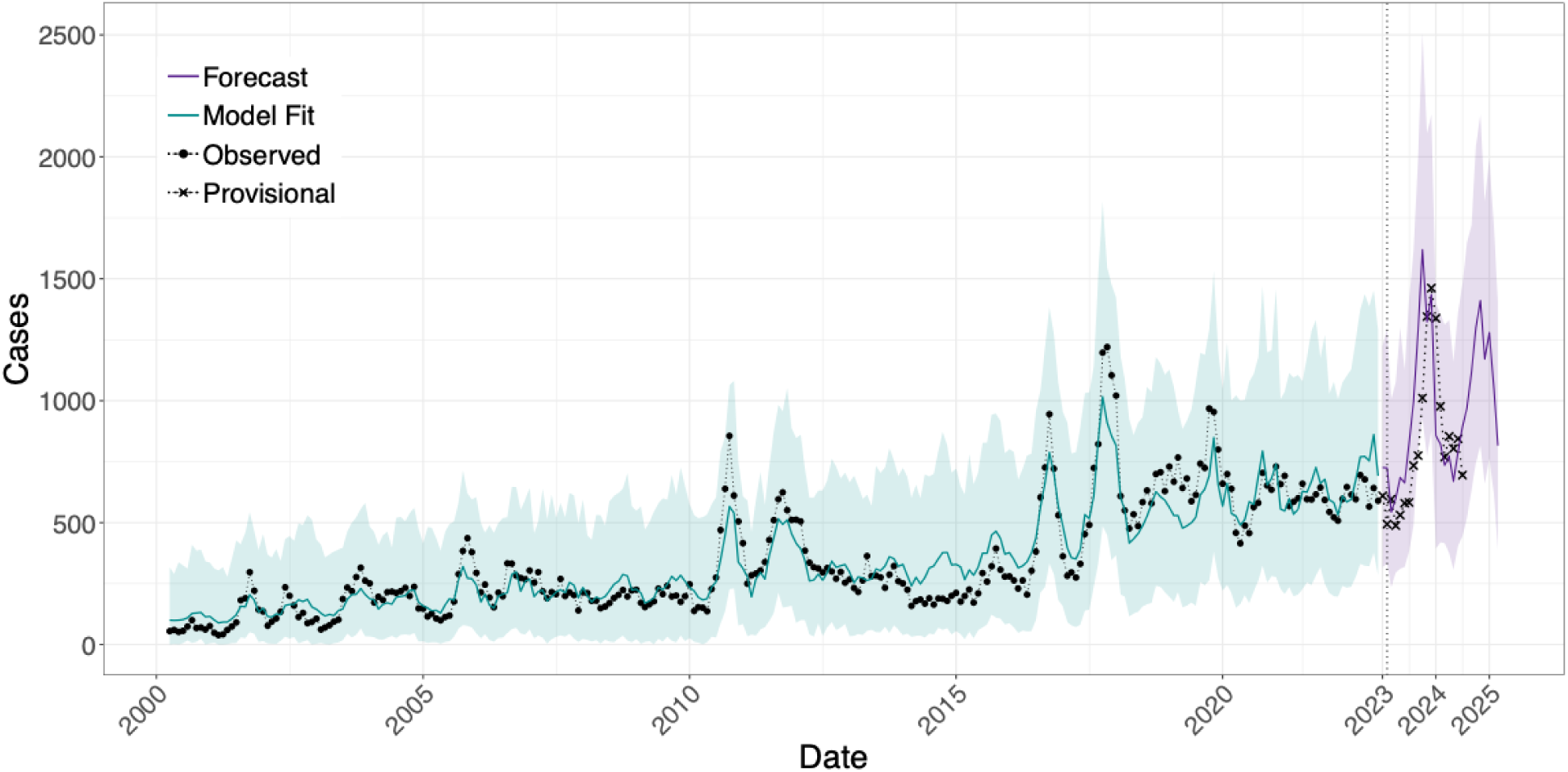
Statewide monthly coccidioidomycosis cases, 2000–2024. Black dots indicate confirmed cases reported between 2000–2022, x’s indicate the provisional cases reported between Jan. 1, 2023, and Jul. 31, 2024, the green line represents the ensemble model fit to the observed case data (R^2^ = 0.87), and the purple line indicates the ensemble model predicted (Apr. 1, 2023 – Mar. 31, 2024) and forecasted (Apr. 1, 2024 – Mar. 31, 2025) cases between Apr. 1, 2023, and Mar. 31, 2025. 90% prediction intervals are represented by the colored bands.

**Fig. 3.**
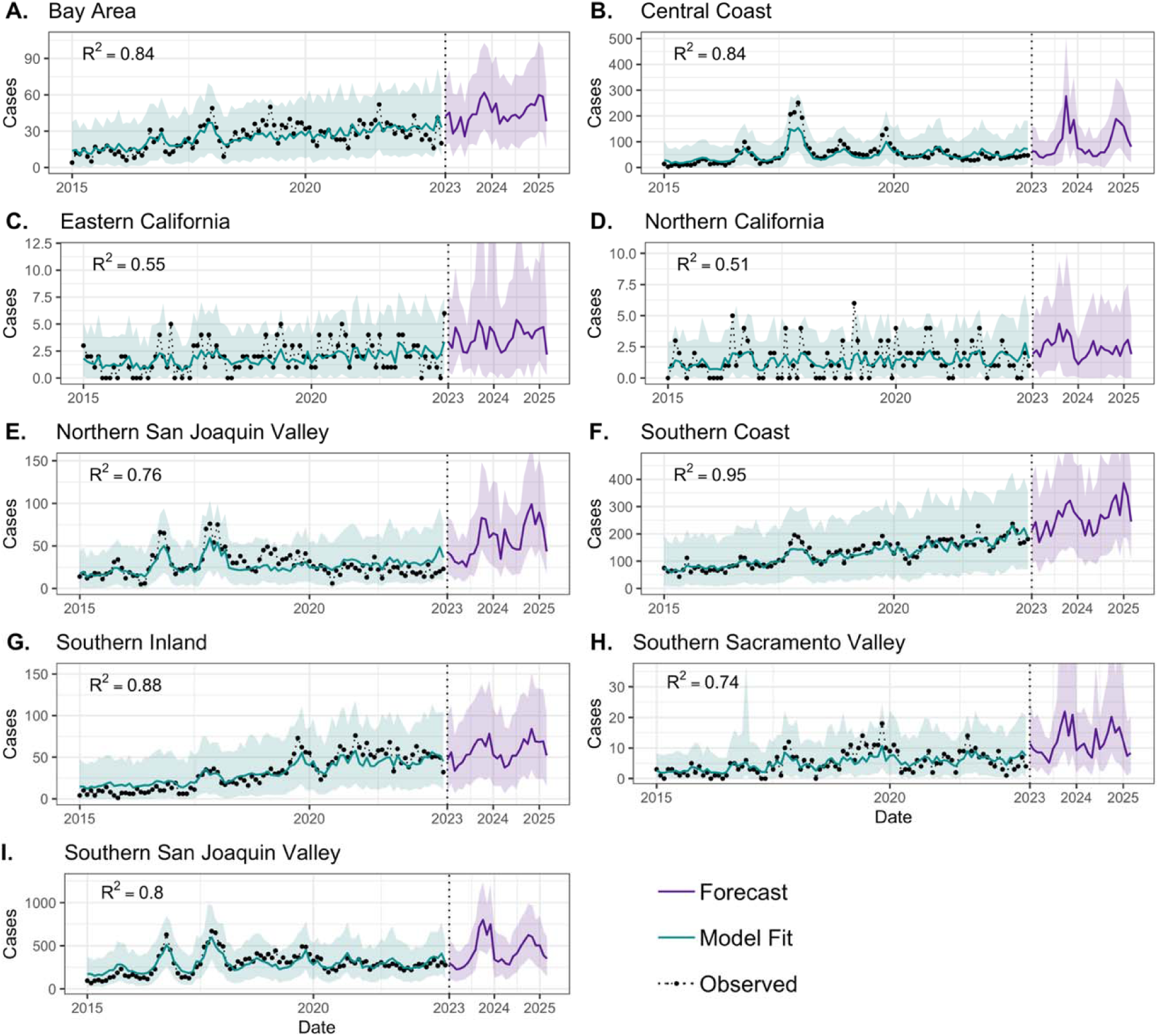
Regional monthly coccidioidomycosis cases, 2015–2024. Black dots indicate confirmed cases reported between 2015–2022, the green line represents the ensemble model fit to the observed case data, and the purple line indicates the ensemble model predicted (Apr. 1, 2023 – Mar. 31, 2024) and forecasted (Apr. 1, 2024 – Mar. 31, 2025) cases between Jan. 1, 2023, and Mar. 31, 2025. 90% prediction intervals are represented by the colored bands.

**Table 1.** Region-level forecasted incident cases for the 2023-2024 (Apr. 2023–Mar. 2024) and 2024-2025 (Apr. 2024–Mar. 2025) transmission years. PI = prediction interval.

| Region | 2023-2024 Predicted (90% PI) | 2024-2025 Forecasted (90% PI) |
| --- | --- | --- |
| Bay Area | 529 (427–665) | 558 (441–675) |
| Central Coast | 1,189 (819–1,599) | 1,207 (867–1,585) |
| Eastern California | 42 (27–81) | 49 (27–84) |
| Northern California | 32 (19–45) | 28 (15–43) |
| Northern San Joaquin Valley | 605 (474–801) | 811 (646–1,044) |
| Southern Coast | 3,049 (2,495–3,739) | 3,322 (2,694–3,961) |
| Southern Inland | 694 (549–902) | 725 (537–919) |
| Southern San Joaquin Valley | 5,557 (4,778–6,757) | 5,399 (4,556–6,442) |
| Southern Sacramento Valley | 149 (108–516) | 147 (107–282) |
| **Statewide** | **11,846 (10,056–14,094)** | **12,244 (10,476–14,506)** |

### Coccidioidomycosis cases forecasted to remain high in 2024-2025 season

Our model forecasts a similarly high incidence between April 1, 2024, and March 31, 2025, forecasting 12,244 cases (90% PI: 10,476–14,506) to occur statewide — a 62% increase over the transmission year two years prior. Similar to 2023, the Southern San Joaquin Valley (5,398 cases, 90% PI: 4,556–6,442), Southern Coast (3,322, 90% PI: 2,694–3,961), and Central Coast (1,207 cases, 90% PI: 867–1,585) regions are expected to see the largest number of infections (Fig. 3; Table 1). Our model forecasts that disease incidence will exhibit pronounced seasonality, particularly in endemic regions (Fig. 3), with cases rising in June and peaking in November at 1,411 (90% PI: 815–2,172) cases statewide – 98% higher than the peak in 2022 (714) and nearly as high as the peak in 2023 (1,462) (3). When assuming that precipitation patterns in the remainder of 2024 and into 2025 would be drier-than-average (20^th^ percentile of rainfall) vs. wetter-than-average (80^th^ percentile), our model forecasts 12,317 and 11,866 cases, respectively, suggesting that precipitation that has already occurred is a larger driver of incidence than concurrent rainfall (Fig. 4). Similarly, when assuming the temperature will be 3°F above or below average, our model forecasts 12,652 and 12,286 cases, respectively (Fig. 4).

**Fig. 4.**
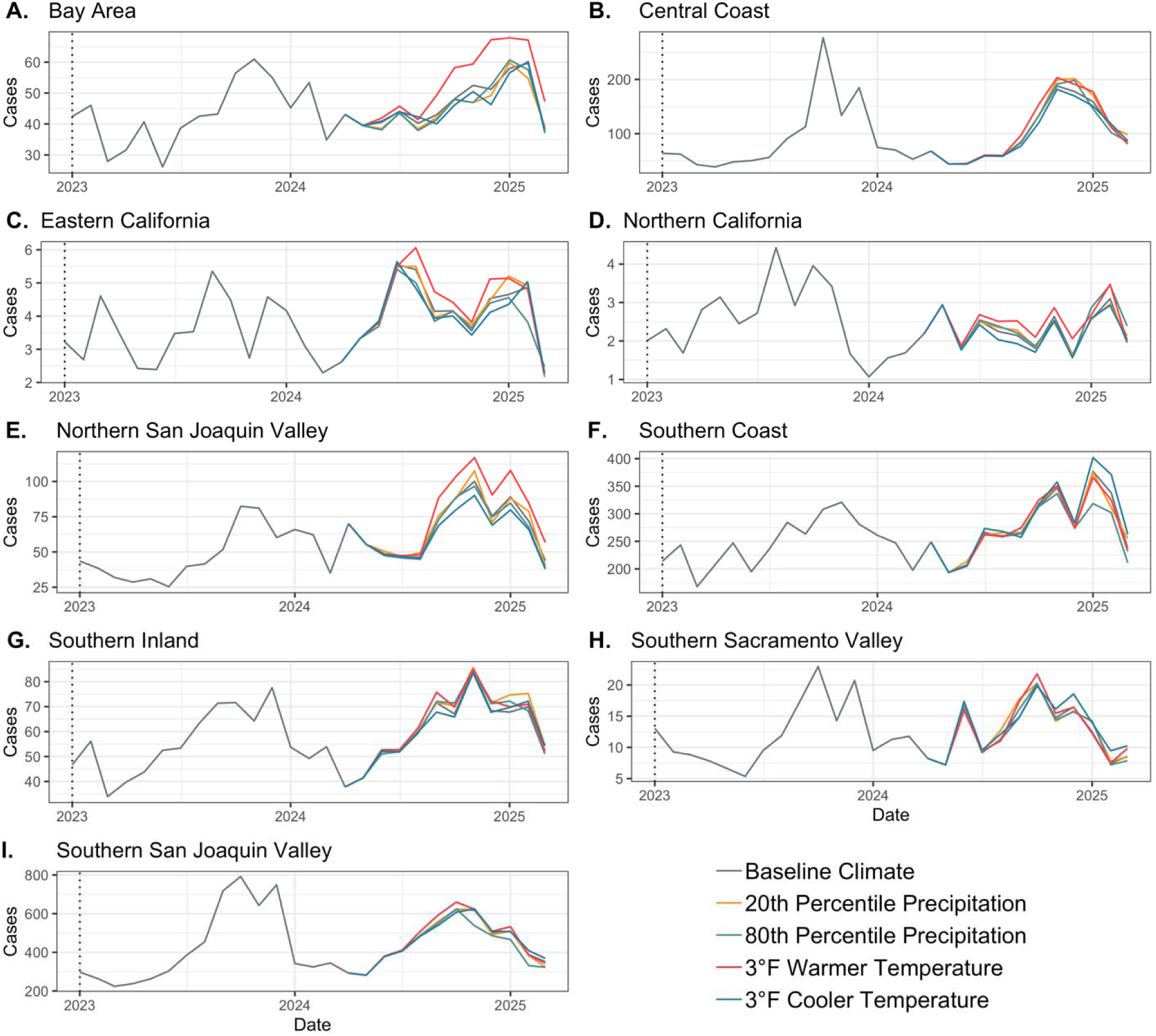
Forecasted regional monthly coccidioidomycosis cases between January 1, 2023, and March 31, 2025, under varying future climates in 2024-2025. The baseline climate scenario represents the 50^th^ percentile of precipitation between 1981–2023 and extrapolated monthly average temperatures assuming a 42-year linear trend. The 20^th^ and 80^th^ percentile precipitation scenarios assume the baseline temperature scenario, and the 3°F warmer or cooler temperature scenarios assume the baseline precipitation scenario (i.e., 50^th^ percentile).

## Discussion

Our outlook for 2024-2025 suggests another high-risk year for coccidioidomycosis in California and predicts a seasonal peak in reported incidence during the fall months. Climate change will likely bring profound changes in California’s hydroclimate, with some of these shifts already underway (12, 13). While trends in mean annual average precipitation in California are likely to be modest (14), precipitation variability will likely increase considerably (15), including increasingly large and frequent swings between high and low precipitation conditions both from season to season and between years—a phenomena known as “precipitation whiplash” (13, 16). Warming temperatures are directly driving increasing rates of evaporative demand (17) and soil drying (18), ultimately increasing the frequency, severity, and rapidity of onset of soil moisture droughts in California (12). Together, the observed and projected shifts in California’s precipitation characteristics and the amplified drying effects of rising temperatures will likely give rise to increasingly pronounced swings between dry and wet soil moisture conditions than might arise from either factor in isolation. Given recent patterns in coccidioidomycosis incidence and their association with transitions from anomalously dry to wet years, increasing swings may broadly increase coccidioidomycosis risk across California as well as adjacent regions with similar hydroclimate dynamics.

The dry summer and fall months are likely to be the highest risk for *Coccidioides* exposure, as drier soil facilitates dust and the release of *Coccidioides* spores. Reported cases typically lag pathogen exposure by 1-2 months, aligning with observed peak of cases around November (3, 19). To prevent exposure, Californians living and working in the endemic and emerging regions should avoid common sources of dust as much as possible, practice dust suppression techniques, and consider using N95 masks when disturbing soil (e.g., in construction). Examples of dust suppression techniques include limiting vehicle speed through dusty roads, installing hoods and filters where possible to vent dusty processes, and installing fencing, windbreaks, and vegetation where possible, among others (20). Furthermore, clinicians should consider coccidioidomycosis as a potential diagnosis when evaluating a patient presenting with community acquired pneumonia or a respiratory illness (i.e., symptoms such as cough, fever, fatigue, and difficult breathing) who lives in, works in, or travelled to an endemic or emerging region, particularly patients who do not respond to an initial course of antibiotics, who had significant exposure to dust or dirt, or whose symptoms last more than 1-2 weeks.

The findings in this report are subject to at least three limitations. First, the analysis is subject to exposure misclassification as case data were aggregated to month of estimated disease onset, which tends to lag the month of *Coccidioides* exposure by 1-2 months, and census tract of case residence, which may not represent the true location of exposure (e.g., occupational exposures). Second, climate conditions in the forecasted period are uncertain and variation in future climate may impact disease incidence; however, forecasts were similar under the several alternate climate scenarios examined. Third, our forecasts cannot account for stochastic point source outbreaks that may lead to anomalously high case counts in certain regions. Finally, the publishing of this report may impact the accuracy of the forecasts by, for example, increasing case suspicion among physicians or influencing risk reduction behaviors among the public.

## Conclusions

Alternation between dry and wet years has been linked to increased coccidioidomycosis in California. A particularly wet 2022-2023 proceeded by extreme drought in 2020-2022 may have been responsible for near record case counts observed in 2023 in California. We develop an ensemble model that generated an accurate prediction of high cases in 2023 and forecasts a similarly high transmission season for 2024-2025. Predictive models that forecast disease risk, such as those developed here, can guide the creation and dissemination of state and local public health alerts by identifying when and where disease risk is expected to be highest. Raising provider and patient awareness about disease risk and presentation may lead to earlier diagnosis and improved disease outcomes.

## Data Availability

The coccidioidomycosis surveillance data used here is not publicly available and is only available with explicit permission from the California Department of Public Health.

## Acknowledgements

Research reported in this manuscript was supported by the National Institute of Allergy and Infectious Diseases (NIAID) of the National Institutes of Health under award numbers R01AI148336, R01AI176770, and K01AI173529. The content is solely the responsibility of the authors and does not necessarily represent the official views of the National Institutes of Health.

## Supplemental Tables and Figures

**Fig. S1.**
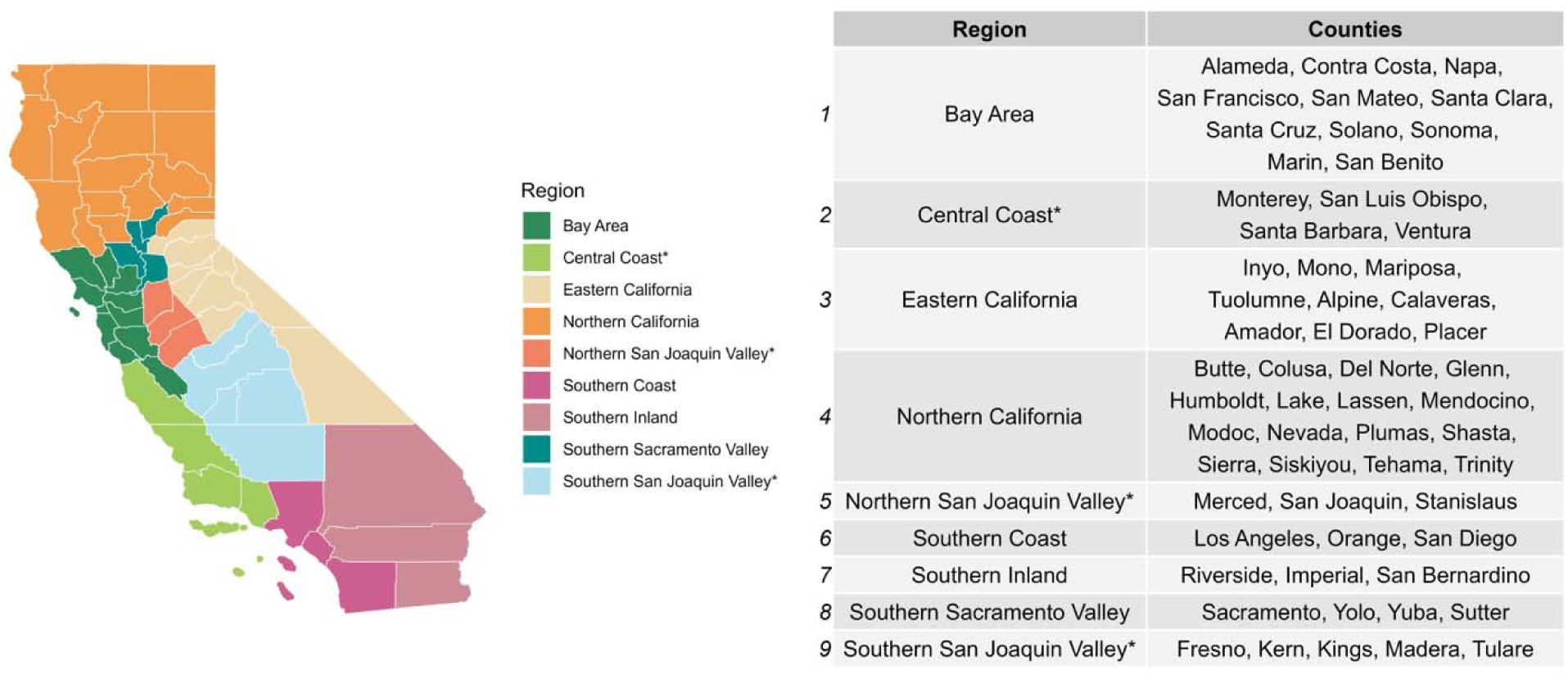
Map of California counties by region. Starred regions are considered “high incidence”; counties within these regions were modelled independently.

**Fig. S2.**
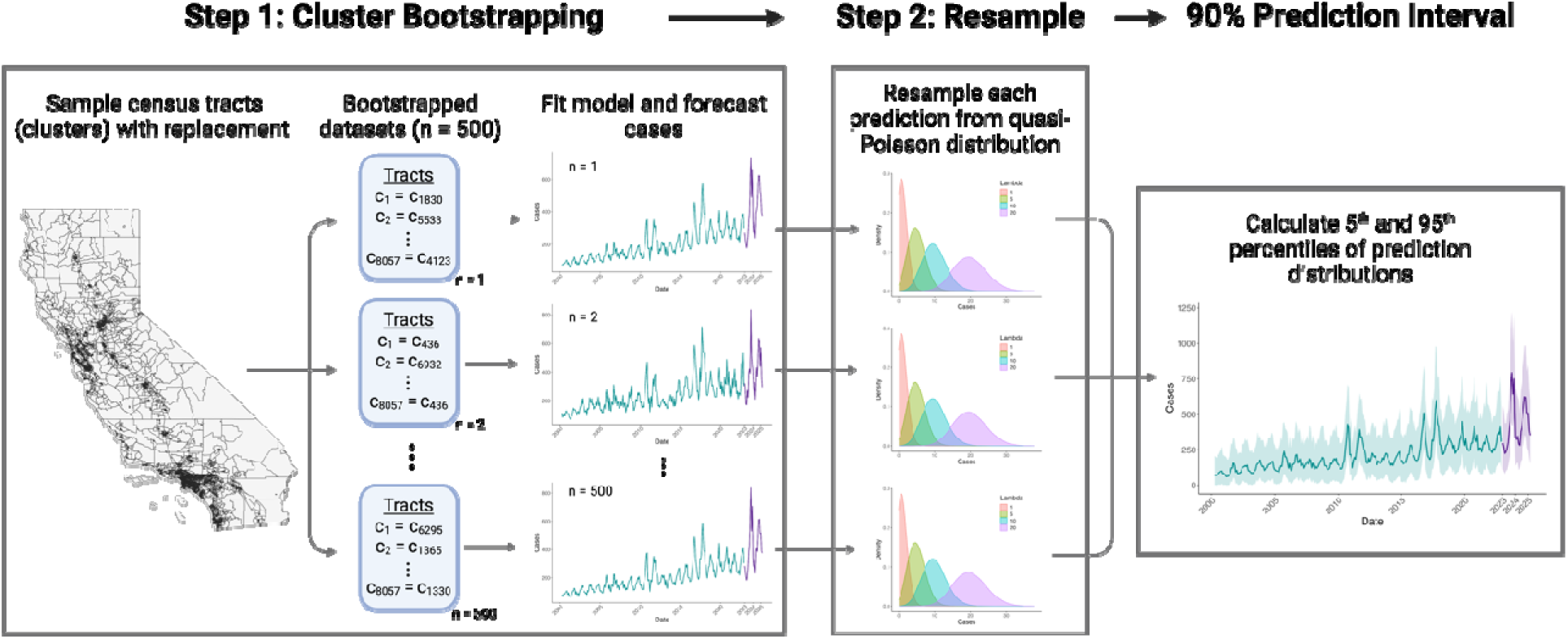
To generate 90% prediction intervals, we used a two-step bootstrapping process. In step 1, we sampled census tracts (n = 8,057) with replacement to generate 500 datasets, fit the models to each dataset (n = 1–500), and forecasted cases using each model. In step 2, we then resampled each estimate from a quasi-Poisson distribution with the mean set to each bootstrapped prediction and calculated the 5^th^ and 95^th^ percentile of the resulting distribution to obtain the 90% prediction interval. Figure created with BioRender.

**Table S1.**
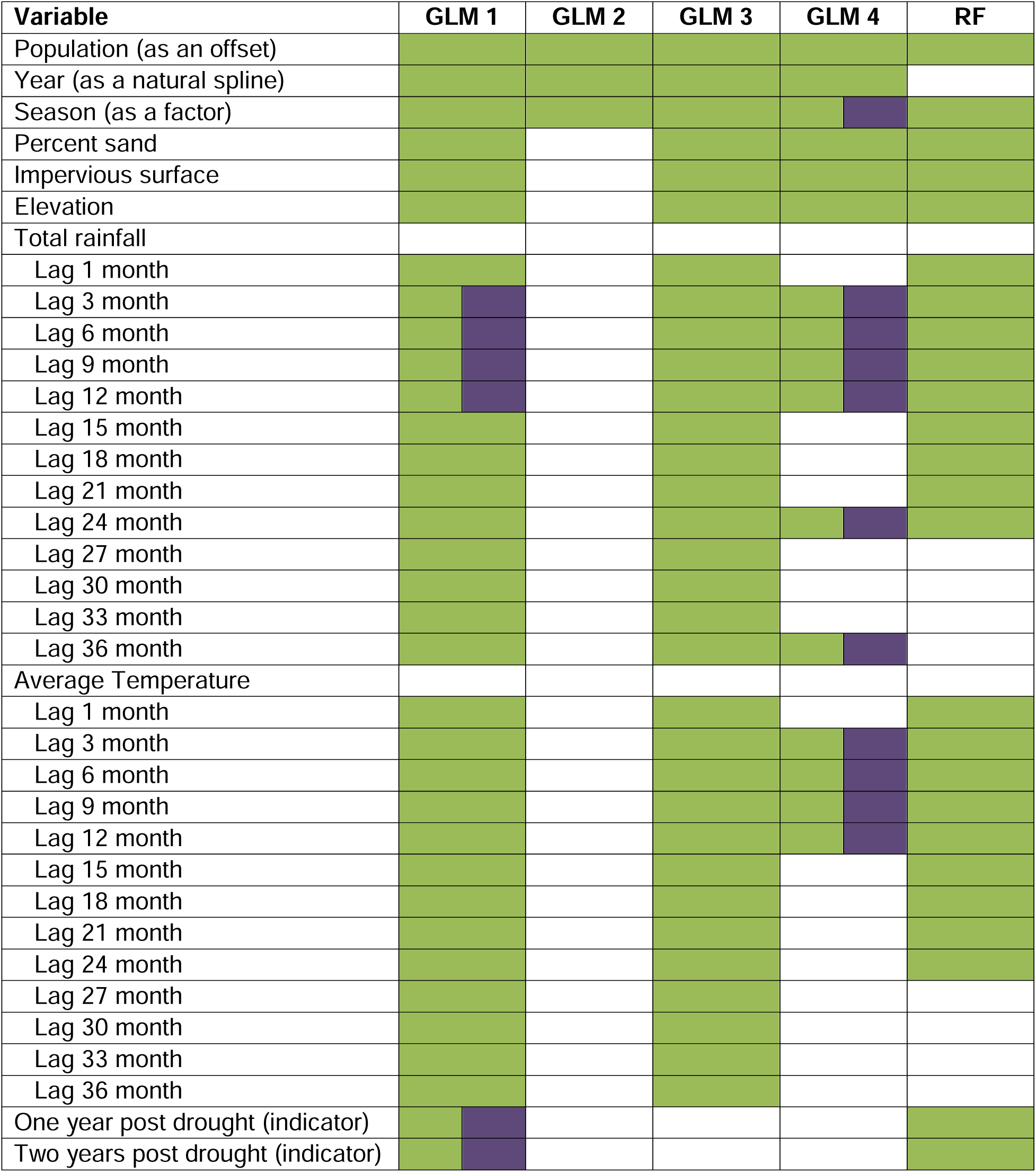
Variables included in the five models included in the ensemble. Green cells were used as main effects and purple cells were used as interactions. Models 1-4 were generalized linear models (GLMs) and model 5 was a random forest (RF).

